# Empirical genomic methods for tracking plasmid spread among healthcare-associated bacteria

**DOI:** 10.1101/2022.09.09.22279653

**Authors:** Daniel Evans, Alexander Sundermann, Marissa Griffith, Vatsala Srinivasa, Mustapha Mustapha, Jieshi Chen, Artur Dubrawski, Vaughn Cooper, Lee Harrison, Daria Van Tyne

**Affiliations:** Department of Infectious Diseases and Microbiology, University of Pittsburgh Graduate School of Public Health, Pittsburgh PA, USA; Division of Infectious Diseases, University of Pittsburgh School of Medicine, Pittsburgh PA, USA; Center for Genomic Epidemiology, University of Pittsburgh School of Medicine, Pittsburgh PA, USA; Auton Laboratory, Carnegie Mellon University, Pittsburgh PA, USA; Department of Microbiology and Molecular Genetics, University of Pittsburgh School of Medicine, Pittsburgh PA, USA; Center for Evolutionary Biology and Medicine, University of Pittsburgh School of Medicine, Pittsburgh, PA, USA

**Author notes:** Correspondence to Dr. Daria Van Tyne, PhD, Division of Infectious Diseases, University of Pittsburgh School of Medicine, Pittsburgh PA 15213, USA.

## Abstract

**Background:** Healthcare-associated bacterial pathogens frequently carry plasmids that contribute to antibiotic resistance and virulence. The horizontal transfer of plasmids in healthcare settings has been previously documented, but genomic and epidemiologic methods to study this phenomenon remain underdeveloped. The objectives of this study were to develop a method to systematically resolve and track plasmids circulating in a single hospital, and to identify epidemiologic links that indicated likely horizontal plasmid transfer.

**Methods:** We derived empirical thresholds of plasmid sequence similarity from comparisons of plasmids carried by bacterial isolates infecting individual patients over time, or involved in hospital outbreaks. We then applied those metrics to perform a systematic screen of 3,074 genomes of nosocomial bacterial isolates from a single hospital for the presence of 89 plasmids. We also collected and reviewed data from electronic health records for evidence of geotemporal associations between patients infected with bacteria encoding plasmids of interest.

**Findings:** Our analyses determined that 95% of analyzed genomes maintained roughly 95% of their plasmid genetic content at a nucleotide identity at least 99·985%. Applying these similarity thresholds to identify horizontal plasmid transfer identified 45 plasmids circulating among clinical isolates. Ten plasmids met criteria for geotemporal links associated with horizontal transfer. Several plasmids with shared backbones also encoded different additional mobile genetic element content, and these elements were variably present among the sampled clinical isolate genomes.

**Interpretation:** The horizontal transfer of plasmids among nosocomial bacterial pathogens is frequent within hospitals and can be monitored with whole genome sequencing and comparative genomics approaches. These approaches should incorporate both nucleotide identity and reference sequence coverage to study the dynamics of plasmid transfer in the hospital.

**Funding:** This research was supported by the US National Institute of Allergy and Infectious Disease (NIAID) and the University of Pittsburgh School of Medicine.

**RESEARCH IN CONTEXT:** *Evidence before this study:* A search of PubMed for research articles containing the search terms “plasmid”, “transfer”, “epidemiology”, “hospital”, and “patients” identified 115 peer-reviewed manuscripts published before 01 January 2022. Twenty-four manuscripts documented the dissemination of one or more plasmids by horizontal transfer in a hospital setting. Most of these prior studies focused on a single plasmid, outbreak, antibiotic resistance gene or pathogen species, and none established an *a priori* approach to identify plasmids circulating among non-clonal bacterial genomes. While prior studies have quantified plasmid preservation and nucleotide identity, similarity thresholds to infer horizontal transfer were neither uniform across studies nor systematically derived from empirical data.

*Added value of this study:* This study advances the field of genomic epidemiology by proposing and demonstrating the utility of empirically derived thresholds of plasmid sequence similarity for inferring horizontal transfer in healthcare settings. It also advances the field by tracking horizontal plasmid transfer within a single hospital at a hitherto unprecedented scale, examining the evidence of horizontal transfer of 89 plasmids among thousands of clinical bacterial isolates sampled from a single medical center. Our systematic review of patient healthcare data related to horizontal transfer also occurred at a breadth not previously undertaken in hospital epidemiology.

*Implications of all the available evidence:* When successfully integrated into contemporary methods for surveillance of nosocomial pathogens, comparative genomics can be used to track and intervene directly against the dissemination of plasmids that exacerbate virulence and antimicrobial resistance in healthcare-associated bacterial infections. Standardized thresholds of plasmid identity benefit epidemiologic investigations of horizontal transfer similar to those offered by establishing uniform thresholds of genome identity for investigations of bacterial transmission.

## Introduction

Healthcare-associated infections impose a serious burden on healthcare infrastructure and widespread morbidity and mortality, both in the United States and worldwide.^1^ Plasmids carried by bacterial pathogens that cause these infections often carry genes conferring antimicrobial resistance, virulence, and environmental persistence. These features complicate patient care and increase disease severity.^2,3^ While our understanding of plasmid transmission via horizontal transfer among bacteria in healthcare settings has increased in recent years, it remains poorly understood compared to healthcare-associated transmission of bacteria.^4–6^ One substantial gap in the burgeoning field of plasmid epidemiology is the lack of uniform thresholds of sequence similarity by which recent horizontal plasmid transfer can be inferred. Without standardized genomics-based approaches to resolve and characterize plasmids found in healthcare settings, inferences regarding plasmid transmission remain speculative and error-prone. Developing metrics to establish plasmid transmission can aid infection prevention personnel in the spread of multidrug-resistant bacteria in hospitals, helping to intervene directly against the dissemination of antimicrobial resistance and virulence.^2,5,7^

Our study had two objectives. The first was to establish sequence similarity thresholds to infer horizontal transfer of plasmids among bacteria collected from a single hospital. The second was to apply these thresholds to a large dataset of whole-genome sequences of nosocomial bacterial pathogens from a tertiary hospital,^8,9^ and to investigate epidemiologic links between patients infected with genetically unrelated bacteria encoding the same plasmid. The intended outcome was to demonstrate that these methods augment our knowledge of plasmid dynamics in healthcare settings at the scale of an entire hospital, further supporting the value of whole-genome sequencing (WGS) in healthcare epidemiology.^9–11^

## Methods

### Collection of clinical bacterial isolates and corresponding patient data

The study took place at the University of Pittsburgh Medical Center (UPMC) Presbyterian Hospital and the University of Pittsburgh School of Medicine. All bacterial isolates analyzed in this study were collected through the Enhanced Detection System for Hospital-Associated Transmission (EDS-HAT) project, using previously published eligibility criteria for selection of isolates.^8,9,11^ EDS-HAT uses a combination of WGS surveillance of selected hospital-associated pathogens and machine learning of the electronic health record to identify otherwise undetected outbreaks and the responsible transmission routes, respectively. Isolates were identified using TheraDoc software (Version 4.6, Premier, Inc, Charlotte NC). The University of Pittsburgh Institutional Review Board (IRB) approved the EDS-HAT project and classified it as being exempt from informed consent because direct contact with human subjects is not performed. Approval was also granted (STUDY20060252) to use data collected for the EDS-HAT project to study the horizontal transfer of mobile genetic elements in the hospital.

### Short- and long-read whole-genome sequencing, processing, and assembly

DNA was extracted from overnight cultures of single bacterial colonies using a Qiagen DNeasy Blood and Tissue Kit (Qiagen, Germantown MD). Short-read WGS was performed using the Illumina platform (Illumina, San Diego CA), with libraries constructed with a Nextera DNA Sample Prep Kit with 150bp paired-end reads and sequenced on the NextSeq platform.

Trim Galore v0.6.1 was used to remove sequencing adaptors, low-quality bases, and poor-quality reads.^12^ Bacterial species were identified from processed Illumina reads by alignment to Kraken v1.0 and RefSeq databases.^13,14^ Genomes of strains sequenced only with Illumina technology were assembled using SPAdes v3.11.^15^ The quality of assembled genomes was then verified using QUAST.^16^ Assembled genomes were excluded if they failed to meet the following quality control parameters: genome-wide read depth of at least 40X, cumulative length of assembled genome within 20% of the expected length for the assigned genus, fewer than 400 contigs in the assembled genome, and an N50 value of greater than 50,000bp. Illumina sequencing data for all isolates is deposited at NCBI with accession numbers listed in **Appendix 1**.

Long-read sequencing and base-calling of select isolates were performed using Oxford Nanopore technology (Oxford Nanopore Technologies, Oxford, United Kingdom). Libraries were constructed using a rapid multiplex barcoding kit (catalog number SQK-RBK004).

Sequencing was performed using an Oxford Nanopore MinION device with R9.4.1 flow cells. Base-calling and read processing was performed using Albacore v2.3.3 or Guppy v2.3.1 (Oxford Nanopore Technologies, Oxford, United Kingdom) using default parameters. Hybrid assembly was performed for genomes for which both short- and long-read sequencing data were available and whose short-read only assemblies passed the aforementioned parameters, using Unicycler v0.4.7 or v0.4.8-beta.^17^

### Genome annotation, plasmid assembly, characterization, alignment, and phylogenetic analyses

Assembled genomes and hybrid-assembled plasmids were annotated with Prokka v1.13 or v1.14.^18^ Multi-locus sequence types (STs) were assigned using *mlst* v2.16.1 with PubMLST typing schemes.^19,20^ Antibiotic resistance genes were identified by BLASTn alignments of assembled genomes to the ResFinder v4.1 database.^21,22^ Plasmid replicons were identified by BLASTn alignments of assembled genomes to the PlasmidFinder v2.1 database.^23^ Other genomic features were identified from annotations by Prokka^18^ and by BLASTn alignments to the VFDB database.^24^ Species were defined by grouping isolates with average nucleotide identity (ANI) of at least 95% to one another and less than 95% ANI to genomes of other species.^25^ Whole-genome phylogenies were constructed from core genome alignments generated by Roary v5.18.2^26^ using RAxML v8.0.26 with 1,000 bootstrap iterations.^27^ Plasmid phylogenies were constructed from core genome alignments of reference plasmids made with *snippy-core* v4.4.5^28^ using RAxML v8.0.26 with 1,000 bootstrap iterations.^27^ Annotated plasmids were aligned to one another using EasyFig v2.2.2.^28^ Plasmids were named according to the genus- or species-based identification code of the isolate from which they were first identified and the contig in the hybrid assembly corresponding to the closed plasmid sequence, separated by an underscore.

### Sequence similarity analyses to establish metrics of plasmid transmission

Plasmids were identified from hybrid-assembled genomes of clonal isolates implicated in outbreaks within the same hospital.^10,11^ Contigs were classified as plasmids if they were circularized during hybrid assembly,^17^ were 2-300kb in length, and possessed at least one replicon or conjugative plasmid gene identified by PlasmidFinder, Prokka, or RAST.^18,23,30^ The proportion of plasmid gene content preserved in bacterial genomes (“sequence coverage”) was calculated by searching for plasmid sequences among the contigs of each assembled bacterial genome using BLASTn,^30^ using a nucleotide sequence identity threshold of 95%. Total plasmid nucleotide sequence identity was quantified based on single nucleotide polymorphisms (SNPs) identified by *snippy-core* v4.4.5.^28^

### Resolving plasmids circulating among nosocomial bacterial pathogens in a single hospital

Eighty-nine closed reference plasmid sequences that met aforementioned sequence coverage criteria were selected from whole-genome bacterial sequences constructed using previously published methods (**Appendix 2**).^5,9,11^ Plasmids were de-duplicated by aligning and visualizing closed reference plasmid sequences of similar lengths and distribution among STs and species using Mauve or EasyFig v2.2.2.^29,31^ 3,074 whole-genome sequences from clinical bacterial isolates were then screened for the presence of 89 plasmids by calculating sequence coverage by BLASTn-based contig mapping^32^ and sequence identity using *snippy-core*,^28^ as described above (**Supplementary Figure 1)**. Plasmids were considered potentially involved in vertical transmission and/or horizontal transfer if they were present at sufficient nucleotide similarity and sequence coverage in at least two bacterial genomes of the same or different bacterial species.

### Identification of geotemporal associations where horizontal plasmid transfer may have occurred

Geotemporal associations between plasmid-carrying isolates were identified by systematic review of patient electronic health record (EHR) data, using a previously published machine learning algorithm based on case-control methodology followed by manual review of the algorithm’s findings.^33^ Geotemporal links highlighted by the algorithm were manually evaluated if (1) they included bacterial isolates of different STs, species, or genera; (2) at least one patient was infected during, after, or shortly before the date of culture of their plasmid-carrying isolate while in the location; and (3) at least one other patient later infected by another isolate with the same plasmid was exposed to the same location within 90 days of the date(s) of culture of their isolate(s). Admission records for roommates with overlapping stays were identified as shared exposures regardless of the length of cohabitation. Manual investigations of patient clusters of interest were performed by a board-certified infection preventionist (AJS), with the investigator blinded to genomic data regarding associated pathogens or plasmids.

## Results

### Empirical thresholds of plasmid similarity to indicate potential horizontal transfer

While the epidemiology of plasmid transfer among hospital-associated bacteria has previously been described,^4,5,34,35^ the field lacks consistent guidelines for determining when horizontal plasmid transfer among infection-derived bacterial isolates can be inferred. We hypothesized that a nucleotide sequence similarity-based threshold could be developed by first quantifying the sequence similarity of plasmids that were carried by bacterial strains known to have been transmitted among hospital inpatients. We first examined four plasmids carried by three clusters of carbapenemase-producing *Klebsiella pneumoniae* and vancomycin-resistant *Enterococcus faecium* that showed strong genomic and epidemiologic evidence of nosocomial transmission within our hospital.^10,11^ We also examined plasmids present in pairs or groups of clinical isolates of identical species and sequence types that were isolated from the same patients at different dates during their hospital stays. These plasmids were presumed to be vertically transmitted during bacterial cell division and long-term carriage within each patient. In total, we performed 57 comparisons between 25 plasmids in 47 bacterial genomes; 15 of these comparisons were between same-patient isolates. Elapsed time between dates of culture of plasmid-linked strains ranged from zero days to 427 days (mean 119.5 days, median 82 days).

We next used a previously published BLASTn-based method to calculate the proportion of each plasmid’s sequence that was preserved in linked isolates.^5^ We also quantified the number of single nucleotide polymorphisms (SNPs) among the shared plasmid sequences present in each group of epidemiologically linked isolates. We refer to these metrics as “plasmid sequence coverage” and “plasmid sequence identity”, respectively **(Figure 1a)**. Ninety-five percent of these comparisons had sequence coverage values of at least 93·7% **(Figure 1b)** and nucleotide sequence identity of at least 99·985% (i.e. fewer than 15 SNPs per 100kb of plasmid sequence, or1 SNP per 6·67kb) **(Figure 1c)**. These results provided initial estimates of sequence similarity thresholds that were then applied for downstream analyses to detect horizontal transfer within the hospital environment.

**Figure 1:**
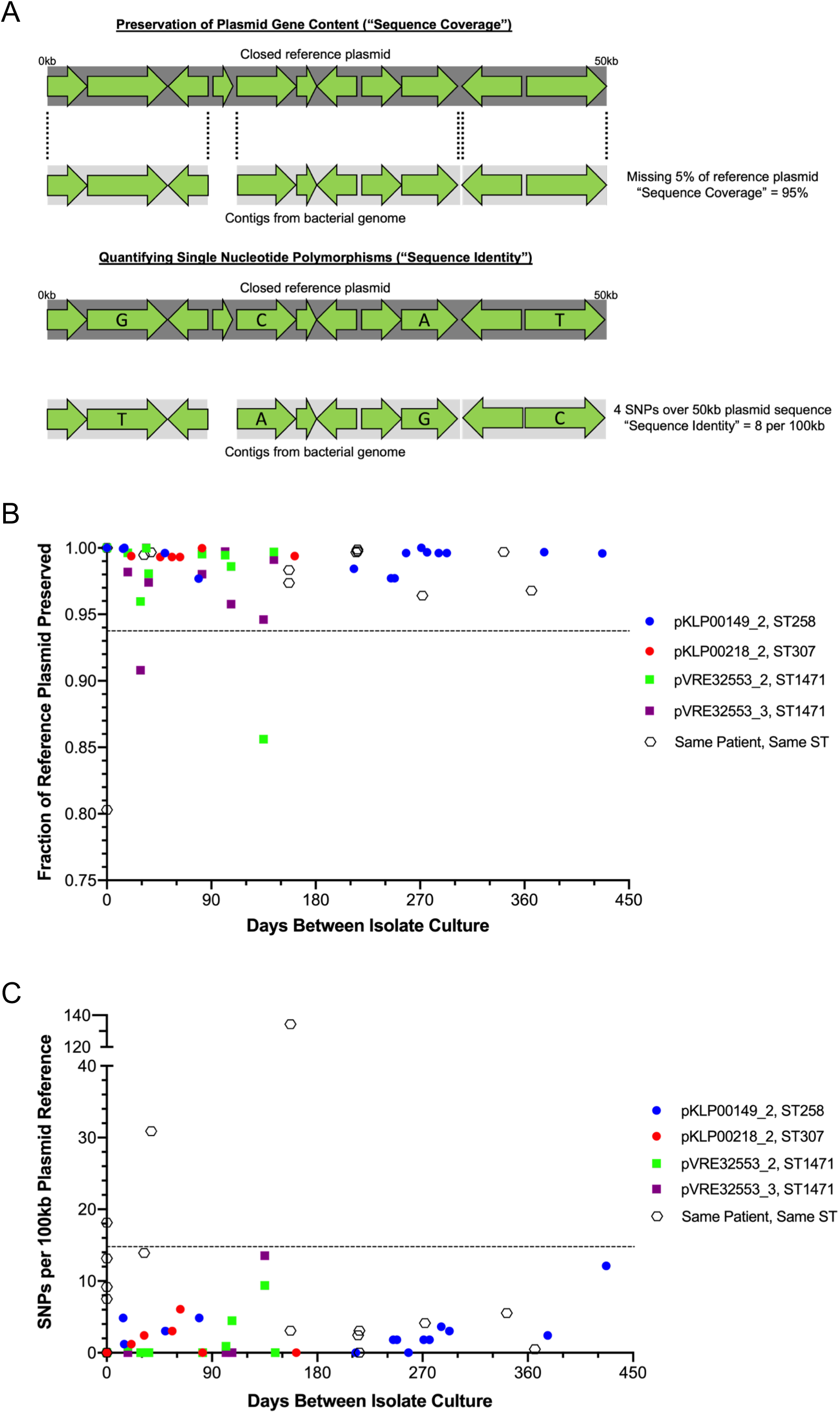
Empirically derived metrics of plasmid sequence similarity to infer horizontal plasmid transfer. (A) Approaches used to calculate (top) plasmid sequence coverage and (bottom) plasmid sequence identity. (B) Sequence coverage of plasmids carried by bacteria engaged in transmission between patients or prolonged carriage in the same patient. Dotted line shows the 5th percentile of coverage among analyzed plasmids. (C) Sequence identity of plasmids carried by isolates engaged in transmission between patients or prolonged carriage in the same patient. Dotted line shows the 95th percentile of SNPs per 100kb of plasmid reference sequence.

### Systematic identification of plasmid horizontal transfer from a single hospital

To systematically identify potential horizontal transfer of plasmids within our hospital, we applied our empirical thresholds described above to search for the presence of 89 reference plasmids in the genomes of 3,074 clinical bacterial isolates collected from 2,086 patients **(Supplementary Figure 1) (Appendix 2)**. Twelve of these plasmids were previously described in a prior study of mobile genetic element diversity in our hospital.^5^ We identified and additional 34 plasmids carried by isolates from more than one patient that had sequence coverage ≥95% and sequence identity ≥99·985% compared to the reference plasmid sequence. The 46 shared plasmids were present in 336 of 3,074 (10·9%) bacterial isolates and in seven of 12 (58·3%) genera in our genomic dataset **(Table 1)**. Bacterial isolates carrying shared plasmids were collected from 291 of 2,086 (14%) unique patients. Shared plasmids were most abundant among isolates belonging to *Enterococcus* (19 plasmids in 214 isolates, 85·6% of *Enterococcus* isolates in the dataset) (**Figure 2a**) and *Klebsiella* (16 plasmids in 77 isolates, 56·6%) **(Figure 2b)**. Nine of the 46 shared plasmids were detected in isolates belonging to different species, and 23 others were found in isolates of different multi-locus STs within the same species, suggesting horizontal plasmid transfer (**Appendix 2**). The remaining 15 plasmids were found in isolates belonging to the same species and ST, suggesting they may have been vertically transmitted between patients along with the bacteria carrying them. Taken together, these results demonstrate the widespread distribution of numerous plasmids engaged in vertical transmission and horizontal transfer within a single hospital.

**Figure 2:**
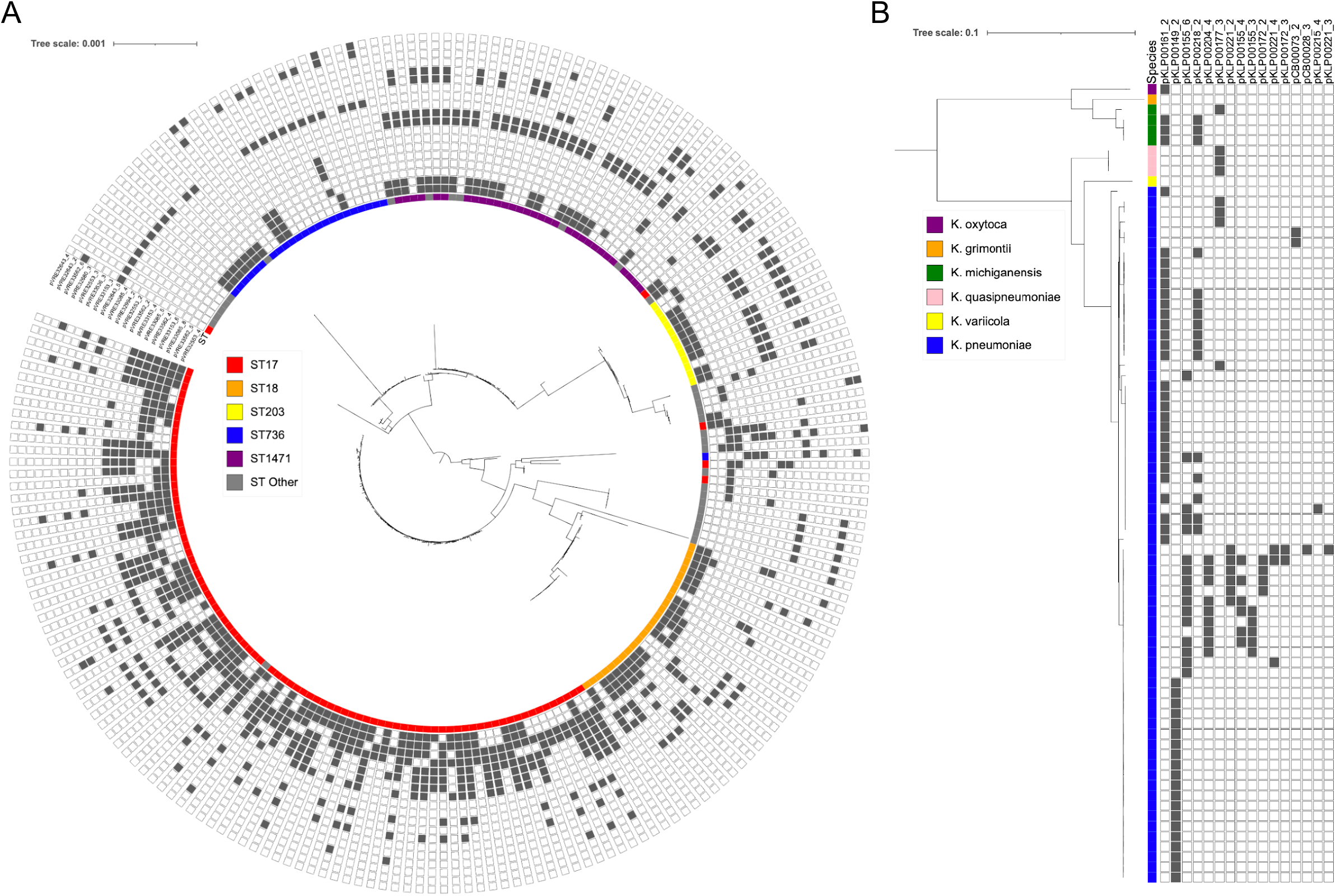
Plasmid distribution among nosocomial pathogens. Bootstrapped RAxML phylogenies of (A) vancomycin-resistant *Enterococcus faecium* and (B) *Klebsiella* spp. carrying shared plasmids. Color strips denote (A) sequence type (ST) of *E. faecium* or (B) species of *Klebsiella*.

### Geotemporal associations among patients implicated in horizontal plasmid transfer

To investigate the epidemiology of horizontal plasmid transfer, we applied a previously published screening algorithm^33^ to systematically review hospital electronic health record (EHR) data of patients infected with isolates belonging to different STs, species, or genera that shared plasmids with high nucleotide identity and high sequence coverage. Using this approach, we identified 18 groups of patients with geotemporal associations who were infected with isolates encoding eight different plasmids that were likely horizontally transferred **(Table 2, Appendix 2)**. These associations fell into three major categories, in which previously uninfected patients were found to be culture-positive for plasmid-carrying pathogens after they were: (1) admitted to rooms previously occupied by infected patients, (2) admitted to units simultaneously occupied by at least one infected patient, or (3) admitted to units previously occupied by at least one infected patient. These findings suggest that plasmids are frequently horizontally transferred among pathogens infecting hospitalized patients independent from bacterial transmission.

### Nucleotide sequence identity threshold improves identification of horizontally transferred plasmids

To investigate whether the methods we developed provided improved resolution over our previously published sequence coverage-based approach,^5^ we reanalyzed ten plasmids that were resolved and investigated in our earlier study. We found that a small ColRNAI plasmid (pKLP00155_6, 9·5kb) that was detected in a large number of isolates of multiple species was most likely two closely related but distinct plasmids that were co-circulating in our hospital **(Figure 3a**). Pairwise comparisons to the pKLP00155_6 reference sequence revealed a group of 13 isolates with >95% sequence coverage of the reference plasmid, but whose sequence identities ranged from 42 to 73 SNPs per 100kb. When the same sequences were compared to pCB00073_2, another reference plasmid that had previously been considered the same as pKLP00155_6 based on sequence coverage alone,^5^ their pairwise sequence identities ranged from 0 to 11 SNPs per 100kb. Separation of these two plasmids was further validated by phylogenetic analysis, which clearly delineated the plasmid sequences into two groups **(Figure 3b)**. When we investigated geotemporal associations between patients infected with bacteria carrying these two plasmids (**Table 2**), we found that two patients infected by isolates of different bacterial species carrying pCB00073_2 were both cultured during overlapping stays on the same hospital unit (**Figure 3c**). Unit-based horizontal transmission was also observed for 16 of the 24 patients carrying pKLP00155_6 housed in three different units, with one patient linked to two of these units (**Figure 3d**).

**Figure 3:**
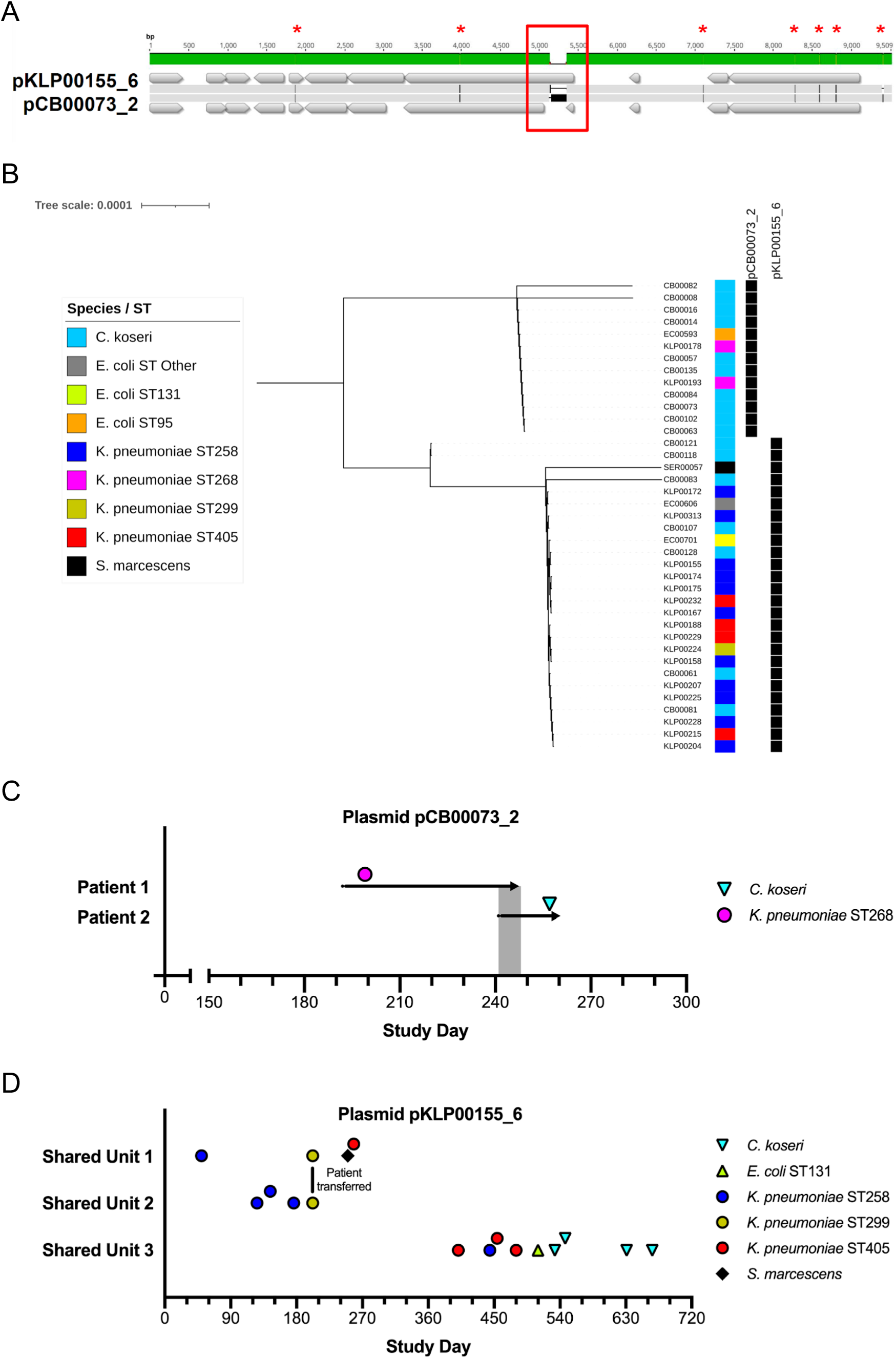
Resolution of two closely related colicin-encoding plasmid variants. (A) Nucleotide sequence alignment with point mutations highlighted by red asterisks and an insertion sequence marked by a red box. (B) Bootstrapped RAxML phylogeny of core plasmid sequences, using the pCB00073_2 sequence as an internal reference. (C) Timeline of dates of culture (de-identified as study days) and inpatient stays (gray shading) of two patients on the same unit infected with bacterial isolates of different species carrying pCB00073_2. (D) Timeline of culture dates of three groups of patients on three different shared units whose nosocomial isolates all carried pKLP00155_6.

We also observed several cases of plasmid pairs that met the sequence identity threshold of ≥99·985% identity, but did not meet the ≥95% sequence coverage threshold. Sequence alignments confirmed that these plasmid pairs each shared a core or “backbone” plasmid sequence, with the longer plasmid of each pair carrying additional genetic cargo of transposases and/or IS6, IS26, and IS110-mediated insertion sequences (**Supplementary Figure 2)**.

Phylogenetic analysis of one of these pairs, pKLP00161_2 and pKLP00218_2, did not separate the plasmids into distinct clades defined by the presence or absence of the additional MGE, suggesting potential gain or loss of plasmid cargo during co-temporal circulation of the bacterial isolates carrying these plasmids (**Supplementary Figure 3**). We considered isolates with high sequence identity and high sequence coverage to one or the other of these plasmid pairs as encoding the same plasmid backbone for the purposes of assessing epidemiologic links between patients.

We next performed extensive reviews of 19 patient care records associated with pathogens of different STs or species that carried four different plasmid variants with shared backbones. Ten patients infected with multiple STs of *K. pneumoniae* and one ST of *K. oxytoca* that all carried the pKLP00218_2 backbone sequence had potential exposures across four different hospital units **(Figure 4a)**. Nine patients infected with five different STs of *E. faecium* carrying the pVRE32553_4 backbone sequence had potential exposures across three different hospital units **(Figure 4b)**. Taken together, these findings demonstrate how empirically derived plasmid similarity thresholds enable tracking of plasmid circulation and horizontal transfer among bacteria. Moreover, these data highlight the potential plasticity of plasmid backbones as they circulate in a hospital.

**Figure 4:**
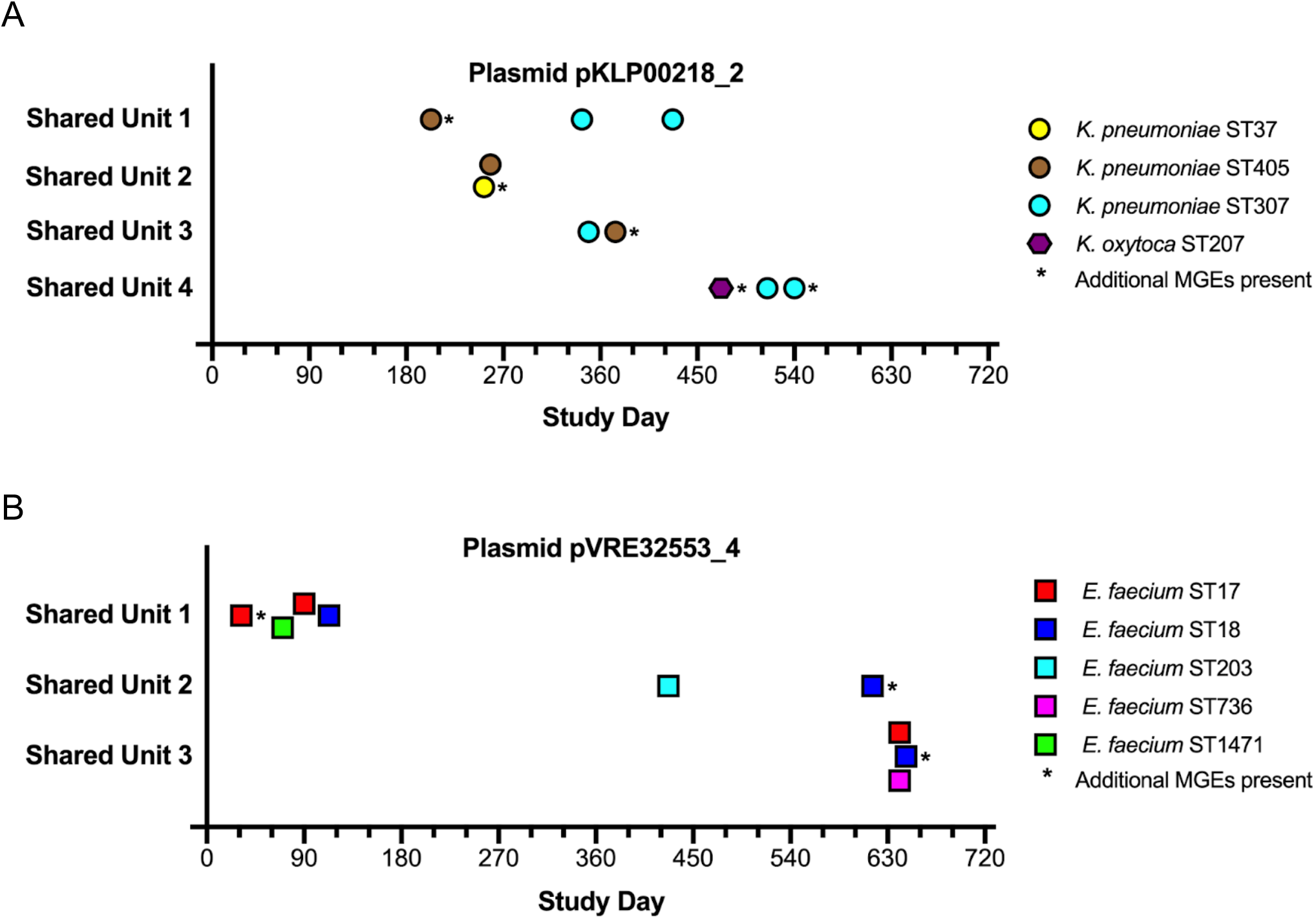
Geographically associated patients infected by bacteria carrying plasmid backbones engaged in horizontal transfer. Plots show associations of patients infected by (A) multiple STs and species of *Klebsiella* spp. carrying plasmid pKLP00218_2, and (B) multiple STs of *E. faecium* carrying pVRE32553_4. Study days correspond to dates of culture of plasmid-carrying pathogens. Shapes and colors of data points correspond to bacterial species and ST, respectively. See Supplementary Figure 2 for plasmid alignments depicting additional mobile genetic element (MGE) sequences.

## DISCUSSION

In this study, we developed uniform thresholds of genetic similarity among plasmids from which horizontal transfer in a hospital might be inferred. We then applied these thresholds to a large dataset of whole-genome sequences of nosocomial bacterial isolates collected from patients at a single medical center. We observed that plasmids potentially engaged in horizontal transfer were widespread among infected patients, and subsequent reviews of electronic health record data found numerous geotemporal links that may have provided opportunities for plasmid transfer between patients. Our findings extend our previous exploration into the genomics of horizontal plasmid transfer within hospital settings,^5^ in which we identified likely plasmid transfer among bacterial pathogens that either co-infected patients or carried those plasmids during transmission between patients with shared geotemporal risk factors.

This study contributes to the field of bacterial genomic epidemiology in several key ways. While numerous methods have been previously applied to resolve and characterize plasmids present among environmental and clinical bacterial isolates from hospitals,^4,5^ these prior studies have largely focused on transmission of antibiotic resistance elements^4,36^ or individual plasmids,^34,35^ often as part of corollary findings in studies focused on bacterial transmission.^6^ In contrast, here we developed and implemented an empirical genome sequence-based approach to systematically cluster plasmids from nosocomial isolates *en masse*. This study also lays the foundation for more detailed investigations into the mechanics of horizontal plasmid transfer within clinical settings^2,35^. Lastly, our work further supports the utility of comprehensive whole-genome sequencing of bacterial isolates from the hospital.^4,5,34^

Our findings bolster several recent insights on the potential causes and effects of plasmid sharing among bacteria infecting hospitalized patients. For example, only 45 of the 89 reference plasmids we studied carried known genes conferring antimicrobial resistance (**Appendix 2**). This finding is in agreement with recent research suggesting that while plasmids mediate much of the spread of antimicrobial resistance, antimicrobial resistance itself may not be the primary driver of horizontal plasmid transfer in clinical settings.^37^ This study also underscores the need for a more thorough characterization of the plasmidome, as the majority of genes on sequenced plasmids have unknown functions.^38^ We also observed widespread sharing of several plasmids among vancomycin-resistant *Enterococcus* (VRE) isolates, corroborating previous findings on the impact of the diversity and extent of plasmid dissemination on enterococcal evolution.^39^

Even with the increased scope of our genomic and epidemiologic analysis, we postulate that our findings still underestimate the true extent to which horizontal plasmid transfer occurs among nosocomial bacterial pathogens. The large number of hybrid-assembled reference plasmids we used to search for evidence of plasmid sharing were sequenced from only 56 isolates from a single hospital,^8^ representing a small subset of the bacteria that infected hospitalized patients during the two-year period of this study. Within this relatively limited dataset, we nonetheless identified several plasmids that exhibited evidence of further recombination and rearrangement while circulating among patients in the hospital. Future studies should further refine the methods we have developed here through analysis of additional reference plasmids as well as isolates sampled from environmental sites or patient colonization. Additionally, computational approaches that resolve plasmids from whole-genome sequence data through non-reference-based approaches^40,41^ could identify additional cases of likely horizontal plasmid transfer.

Our study had several limitations. Our sequence similarity criteria were developed by analyzing a relatively small number of isolates and reference plasmids from a single hospital, which may have introduced selection bias when determining numerical thresholds. Additionally, our dataset of bacterial genomes was subject to selection criteria that limited the sampling of some clinical isolates based on phenotypic antimicrobial resistance profiles and/or body sites.^9,33^ The reference plasmids we studied were largely drawn from sequencing data of select pathogens generated in previous projects,^5,9,42^ rather than from comprehensive efforts that may have yielded more representative samples of plasmids within our hospital. To improve the specificity of our epidemiologic analyses, we limited our scope to geotemporal associations among isolates with substantial taxonomic distance. Lastly, our criteria for inferring horizontal transfer were based on professional expertise in hospital infection prevention and the dynamics of bacterial transmission among hospitalized patients, as knowledge of *in vivo* patterns of plasmid transfer in hospitals is highly limited.^5,35^

In summary, our study advances the field of bacterial genomic epidemiology by applying empirically derived similarity thresholds encompassing both genetic content and nucleotide identity to study plasmid spread among bacterial pathogens. The application of our methods to a large genomic dataset from a single hospital resolved horizontal plasmid transfer at an unprecedented scale and demonstrated the added value of routine whole-genome sequencing of nosocomial bacterial pathogens in hospital infection prevention.^8,9,35^ Applying these refined, plasmid-focused approaches in healthcare settings could help infection prevention teams better understand signs and risk factors for the dissemination of drug resistance, allowing them to improve their approaches to reduce the risk of severe infections among patients.^5,7,35^ Future work in this area should focus on refining similarity thresholds and surveillance methods, further examining patterns of mutation and genetic plasticity among circulating plasmids, and implementing these methods in real-time to control the spread of plasmid-borne antimicrobial resistance and virulence factors that exacerbate nosocomial infections.

## Data Availability

Genome sequence data of all bacterial isolates studied here has been deposited in the Sequence Read Archive (SRA) with accession numbers listed in Appendix 1. Plasmid sequences included in this study have been deposited in GenBank with accession numbers listed in Appendix 2.

https://www.ncbi.nlm.nih.gov/bioproject/PRJNA475751

## ACKNOWLEDGEMENTS

We thank Jane Marsh, Daniel Snyder, Chinelo Ezeonwuka, and Kady Waggle for assistance with clinical isolate collection and whole-genome sequencing. We also thank Melissa Saul for curating electronic health record data for epidemiologic investigations, as well as Alecia Rokes for assistance in curating and classifying reference plasmid sequences. Lastly, we thank Kyle Miller for support in algorithm development and implementation.

---

**Supplementary Figure 1:**
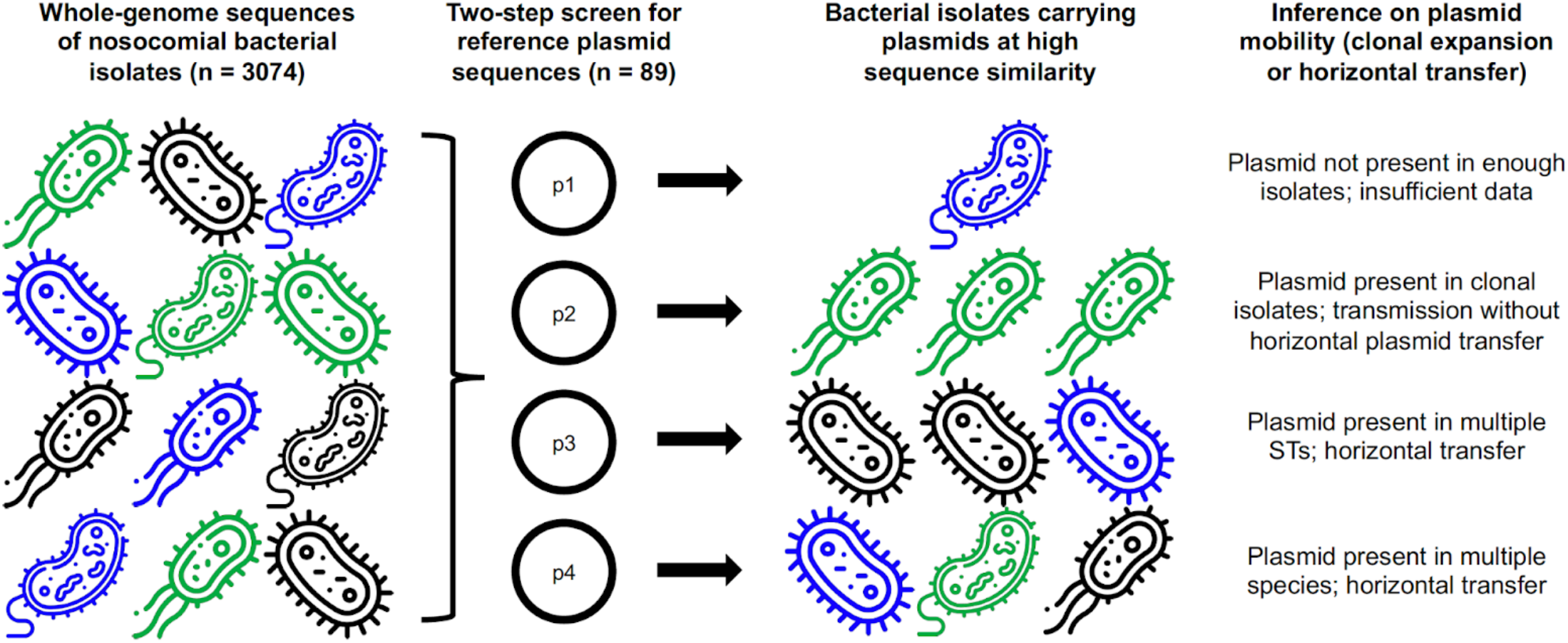
Conceptual flow diagram of alignment-based pipeline to infer horizontal plasmid transfer. Different bacterial icons denote different species, and different colors denote different sequence types (STs). Figure constructed with icons made by Freepik from Flaticon.com.

**Supplementary Figure 2:**
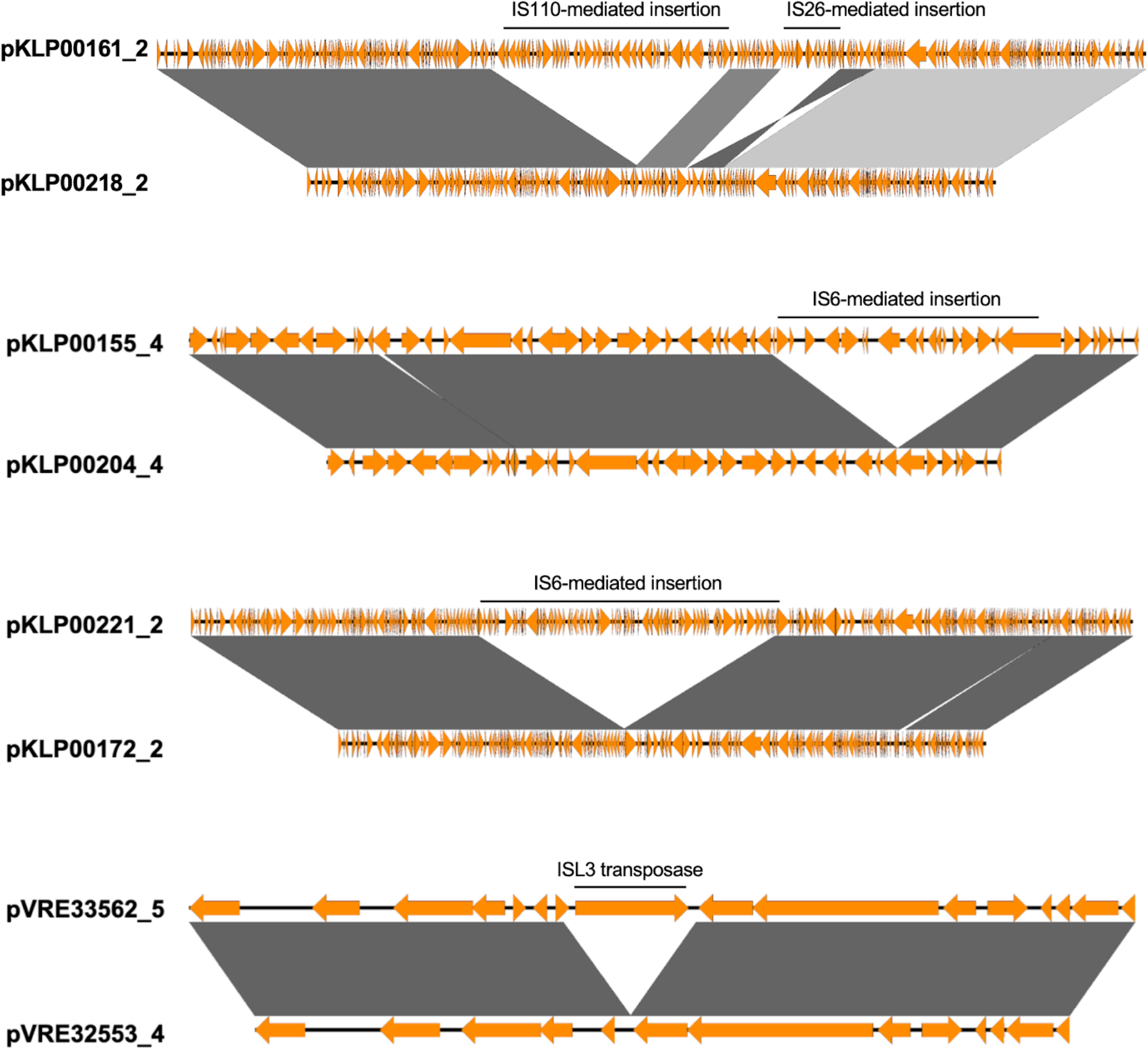
Alignments of four pairs of plasmids with shared backbone sequences that met sequence identity and coverage thresholds. Grey shading indicates a BLASTn alignment of at least 2,000bp with an e-value of 0 at nucleotide sequence identities of at least 99.9%. Darker shading indicates higher nucleotide sequence identity.

**Supplementary Figure 3:**
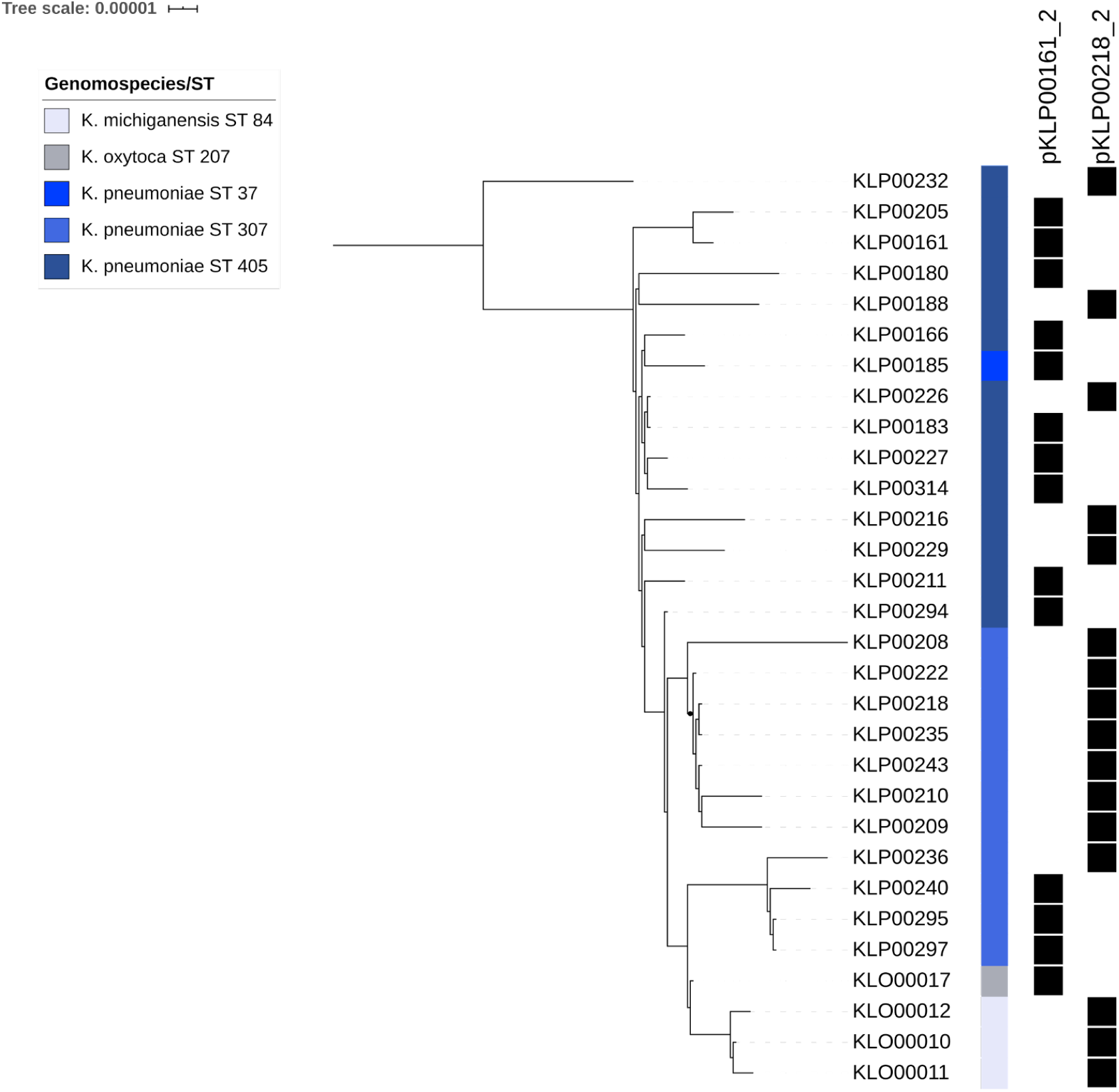
Two recombining variants of *Klebsiella* spp. Plasmids. Bootstrapped RAxML phylogeny of core plasmid sequences conserved among isolates carrying either variant, using the pKLP00218_2 sequence as a reference.

## REFERENCES

1 Centers for Disease Control and Prevention (CDC). 2019 National and State Healthcare-Associated Infections (HAI) Progress Report. 2019. https://arpsp.cdc.gov/profile/national-progress/united-states.

2 Lerminiaux NA, Cameron ADS. Horizontal transfer of antibiotic resistance genes in clinical environments. Can J Microbiol 2019; 65: 34–44.

3 San Millan A. Evolution of Plasmid-Mediated Antibiotic Resistance in the Clinical Context. Trends Microbiol 2018; 26: 978–85.

4 Peter S, Bosio M, Gross C, et al. Tracking of Antibiotic Resistance Transfer and Rapid Plasmid Evolution in a Hospital Setting by Nanopore Sequencing. mSphere 2020; 5: e00525–20, /msphere/5/4/mSphere525-20.atom.

5 Evans DR, Griffith MP, Sundermann AJ, et al. Systematic detection of horizontal gene transfer across genera among multidrug-resistant bacteria in a single hospital. eLife 2020; 9. DOI:10.7554/eLife.53886.

6 Tofteland S, Naseer U, Lislevand JH, Sundsfjord A, Samuelsen O. A long-term low-frequency hospital outbreak of KPC-producing Klebsiella pneumoniae involving Intergenus plasmid diffusion and a persisting environmental reservoir. PloS One 2013; 8: e59015.

7 Salamzade R, Manson AL, Walker BJ, et al. Inter-species geographic signatures for tracing horizontal gene transfer and long-term persistence of carbapenem resistance. Genome Med 2022; 14: 37.

8 Mustapha MM, Srinivasa VR, Griffith MP, et al. Genomic Diversity of Hospital-Acquired Infections Revealed through Prospective Whole-Genome Sequencing-Based Surveillance. mSystems 2022; 7: e01384–21.

9 Sundermann AJ, Chen J, Kumar P, et al. Whole-Genome Sequencing Surveillance and Machine Learning of the Electronic Health Record for Enhanced Healthcare Outbreak Detection. Clin Infect Dis 2021; : ciab946.

10 Sundermann AJ, Babiker A, Marsh JW, et al. Outbreak of Vancomycin-resistant Enterococcus faecium in Interventional Radiology: Detection Through Whole Genome Sequencing-Based Surveillance. Clin Infect Dis Off Publ Infect Dis Soc Am 2019; published online July 16. DOI:10.1093/cid/ciz666.

11 Marsh JW, Mustapha MM, Griffith MP, et al. Evolution of Outbreak-Causing Carbapenem-Resistant Klebsiella pneumoniae ST258 at a Tertiary Care Hospital over 8 Years. mBio 2019; 10. DOI:10.1128/mBio.01945-19.

12 Krueger F, James F, Ewels P, Afyounian E, Schuster-Boeckler B. FelixKrueger/TrimGalore: v0.6.7 - DOI via Zenodo. 2021; published online July 23. DOI:10.5281/ZENODO.5127899.

13 Wood DE, Salzberg SL. Kraken: ultrafast metagenomic sequence classification using exact alignments. Genome Biol 2014; 15: R46.

14 Pruitt KD, Tatusova T, Maglott DR. NCBI reference sequences (RefSeq): a curated non-redundant sequence database of genomes, transcripts and proteins. Nucleic Acids Res 2007; 35: D61–65.

15 Bankevich A, Nurk S, Antipov D, et al. SPAdes: a new genome assembly algorithm and its applications to single-cell sequencing. J Comput Biol J Comput Mol Cell Biol 2012; 19: 455– 77.

16 Gurevich A, Saveliev V, Vyahhi N, Tesler G. QUAST: quality assessment tool for genome assemblies. Bioinforma Oxf Engl 2013; 29: 1072–5.

17 Wick RR, Judd LM, Gorrie CL, Holt KE. Unicycler: Resolving bacterial genome assemblies from short and long sequencing reads. PLoS Comput Biol 2017; 13: e1005595.

18 Seemann T. Prokka: rapid prokaryotic genome annotation. Bioinforma Oxf Engl 2014; 30: 2068–9.

19 Jolley KA, Bray JE, Maiden MCJ. Open-access bacterial population genomics: BIGSdb software, the PubMLST.org website and their applications. Wellcome Open Res 2018; 3: 124.

20 Seemann T. mlst. https://github.com/tseemann/mlst.

21 Bortolaia V, Kaas RS, Ruppe E, et al. ResFinder 4.0 for predictions of phenotypes from genotypes. J Antimicrob Chemother 2020; 75: 3491–500.

22 Zankari E, Hasman H, Cosentino S, et al. Identification of acquired antimicrobial resistance genes. J Antimicrob Chemother 2012; 67: 2640–4.

23 Carattoli A, Zankari E, García-Fernández A, et al. In silico detection and typing of plasmids using PlasmidFinder and plasmid multilocus sequence typing. Antimicrob Agents Chemother 2014; 58: 3895–903.

24 Liu B, Zheng D, Jin Q, Chen L, Yang J. VFDB 2019: a comparative pathogenomic platform with an interactive web interface. Nucleic Acids Res 2019; 47: D687–92.

25 Kim M, Oh H-S, Park S-C, Chun J. Towards a taxonomic coherence between average nucleotide identity and 16S rRNA gene sequence similarity for species demarcation of prokaryotes. Int J Syst Evol Microbiol 2014; 64: 346–51.

26 Page AJ, Cummins CA, Hunt M, et al. Roary: rapid large-scale prokaryote pan genome analysis. Bioinformatics 2015; 31: 3691–3.

27 Stamatakis A. RAxML version 8: a tool for phylogenetic analysis and post-analysis of large phylogenies. Bioinforma Oxf Engl 2014; 30: 1312–3.

28 Seemann, Torsten T. Snippy: rapid haploid variant calling and core SNP phylogeny. Available. 2015. https://github.com/tseemann/snippy.

29 Sullivan MJ, Petty NK, Beatson SA. Easyfig: a genome comparison visualizer. Bioinforma Oxf Engl 2011; 27: 1009–10.

30 Aziz RK, Bartels D, Best AA, et al. The RAST Server: Rapid Annotations using Subsystems Technology. BMC Genomics 2008; 9: 75.

31 Darling ACE. Mauve: Multiple Alignment of Conserved Genomic Sequence With Rearrangements. Genome Res 2004; 14: 1394–403.

32 Altschul SF, Gish W, Miller W, Myers EW, Lipman DJ. Basic local alignment search tool. J Mol Biol 1990; 215: 403–10.

33 Sundermann AJ, Miller JK, Marsh JW, et al. Automated data mining of the electronic health record for investigation of healthcare-associated outbreaks. Infect Control Hosp Epidemiol 2019; 40: 314–9.

34 Prussing C, Snavely EA, Singh N, et al. Nanopore MinION Sequencing Reveals Possible Transfer of blaKPC–2 Plasmid Across Bacterial Species in Two Healthcare Facilities. Front Microbiol 2020; 11: 2007.

35 R-GNOSIS WP5 Study Group, León-Sampedro R, DelaFuente J, et al. Pervasive transmission of a carbapenem resistance plasmid in the gut microbiota of hospitalized patients. Nat Microbiol 2021; 6: 606–16.

36 Cerqueira GC, Earl AM, Ernst CM, et al. Multi-institute analysis of carbapenem resistance reveals remarkable diversity, unexplained mechanisms, and limited clonal outbreaks. Proc Natl Acad Sci U S A 2017; 114: 1135–40.

37 Lopatkin AJ, Huang S, Smith RP, et al. Antibiotics as a selective driver for conjugation dynamics. Nat Microbiol 2016; 1: 16044.

38 Acman M, van Dorp L, Santini JM, Balloux F. Large-scale network analysis captures biological features of bacterial plasmids. Nat Commun 2020; 11: 2452.

39 Arredondo-Alonso S, Top J, McNally A, et al. Plasmids Shaped the Recent Emergence of the Major Nosocomial Pathogen Enterococcus faecium. mBio 2020; 11: e03284-19, /mbio/11/1/mBio.03284-19.atom.

40 Laczny CC, Galata V, Plum A, Posch AE, Keller A. Assessing the heterogeneity of in silico plasmid predictions based on whole-genome-sequenced clinical isolates. Brief Bioinform 2019; 20: 857–65.

41 Robertson J, Nash JHE. MOB-suite: software tools for clustering, reconstruction and typing of plasmids from draft assemblies. Microb Genomics 2018; 4. DOI:10.1099/mgen.0.000206.

42 Babiker A, Evans DR, Griffith MP, et al. Clinical and Genomic Epidemiology of Carbapenem-Nonsusceptible Citrobacter spp. at a Tertiary Health Care Center over 2 Decades. J Clin Microbiol 2020; 58: e00275–20.

